# Global malaria predictors at a localized scale

**DOI:** 10.1101/2023.11.20.23298800

**Authors:** Eloise B. Skinner, Marissa L. Childs, Matthew B. Thomas, Jackie Cook, Eleanore D. Sternberg, Alphonsine A. Koffi, Raphael N’Guessan, Rosine Z. Wolie, Welbeck A. Oumbouke, Ludovic P. Ahoua Alou, Serge Brice, Erin A. Mordecai

## Abstract

Malaria is a life-threatening disease caused by *Plasmodium* parasites transmitted by *Anopheles* mosquitoes. In 2021, more than 247 million cases of malaria were reported worldwide, with an estimated 619,000 deaths. While malaria incidence has decreased globally in recent decades, some public health gains have plateaued, and many endemic hotspots still face high transmission rates. Understanding local drivers of malaria transmission is crucial but challenging due to the complex interactions between climate, entomological and human variables, and land use. This study focuses on highly climatically suitable and endemic areas in Côte d’Ivoire to assess the explanatory power of coarse climatic predictors of malaria transmission at a fine scale. Using data from 40 villages participating in a randomized controlled trial of a household malaria intervention, the study examines the effects of climate variation over time on malaria transmission. Through panel regressions and statistical modeling, the study investigates which variable (temperature, precipitation, or entomological inoculation rate) and its form (linear or unimodal) best explains seasonal malaria transmission and the factors predicting spatial variation in transmission. The results highlight the importance of temperature and rainfall, with quadratic temperature and all precipitation models performing well, but the causal influence of each driver remains unclear due to their strong correlation. Further, an independent, mechanistic temperature-dependent R_0_ model based on laboratory data aligns well with observed malaria incidence rates, emphasizing the significance and predictability of temperature suitability across scales. By contrast, entomological variables, such as entomological inoculation rate, were not strong predictors of human incidence in this context. Finally, the study explores the predictors of spatial variation in malaria, considering land use, intervention, and entomological variables. The findings contribute to a better understanding of malaria transmission dynamics at local scales, aiding in the development of effective control strategies in endemic regions.

## 1 Introduction

Malaria, a vector-borne disease caused by *Plasmodium* parasites, which are transmitted by *Anopheles* mosquitoes, has had a significant impact on human history, influencing our evolution, health, and social structures (Athni et al., 2021). Despite advancements in technology and significant progress in malaria control in the 21st century, it remains a major burden to humanity. In 2021, there were an estimated 247 million malaria cases and 619,000 deaths, with the majority occurring in sub-Saharan Africa (World Health Organization, 2022). Malaria eradication is a long-standing priority for global health and non-profit organizations, who collectively invest over $4.3 billion annually (Feachem et al., 2019). On a global scale, malaria incidence decreased by 37% between 2000 and 2015, and more than half of the world’s countries are now malaria-free (Cibulskis et al., 2016). However, in many malaria hotspots, reductions have not been as significant. In fact, between 2016 and 2017, more than 3.5 million cases of malaria were reported in ten African countries alone (Ryan et al., 2020). Consequently, understanding the local drivers of malaria in endemic hotspots of transmission is critical from both a biological and control perspective.

Malaria transmission is dependent on entomological cycles that drive transmission, and as such, there are many biotic, abiotic, and social variables that can affect incidence. At global and national scales, climate predictors such as temperature and precipitation have been used to model malaria transmission with high success. Mechanistic links between temperature and global malaria transmission are recognised to be nonlinear with transmission constrained between 17°C and 34°C and transmission peaking at 25°C (Carlson et al., 2023; Mordecai et al., 2013; Peterson, 2009; Ryan et al., 2015; Shapiro et al., 2017; Villena et al., 2022; Yamana & Eltahir, 2013) and the most recent estimate being between 19.1°C and 30.1°C (Villena et al., 2022). Rainfall is also recognized to have a nuanced relationship with malaria: accumulated rainfall can create breeding habitats and increase vector abundance, but excessive rainfall can flush out breeding sites and decrease vector abundance (Eikenberry & Gumel, 2018; Paaijmans et al., 2009; Yamana & Eltahir, 2013). Despite the clear mechanistic links, estimating connections between climate and malaria transmission at local scales is challenging because a given location may be below, at, or above optimal temperature and/or can experience high rainfall variability.

Vector indices, including population abundance, species composition, feeding patterns, and entomological inoculation rate (EIR)—which are expected to be directly affected by climate—can provide a more direct estimate of transmission risk. Entomological variables are not as readily available to use in malaria predictions as climate variables because they require detailed surveillance and trained personnel, can be costly, and the resulting data can be highly heterogeneous. Despite this, the use of these variables, and EIR in particular, is considered to be a more direct measure of transmission intensity than vector incidence or prevalence (Kelly-Hope & McKenzie, 2009). It is therefore important to understand how the predictive power of entomological indices compares to that of more easily obtained climate variables for understanding local-scale variation in malaria incidence. Land use, which can influence microclimate, the availability of vector habitat, vector population size, and settings in which vectors bite humans, has also been associated with malaria transmission.

Certain land use practices can create or modify suitable breeding sites for mosquito vectors, leading to increased malaria risk. For example, deforestation and urbanization can create new habitats and increase the proximity of humans to mosquito breeding sites, resulting in higher transmission rates (Afrane et al., 2012; MacDonald & Mordecai, 2019). Additionally, changes in land use patterns can alter microclimates, affecting mosquito abundance and behavior (Afrane et al., 2012; Afrane et al., 2005; Afrane et al., 2006). In sum, coarse and fine scale variation in climate, land use, and human activities combine to determine malaria risk, yet predicting the outcomes of these interacting and nonlinear effects remains a challenge.

Large-scale studies have been informative for developing frameworks to benchmark the success of intervention strategies, to allow for resource allocation, and to identify future populations at risk under a changing climate (Bhatt et al., 2015; Carlson et al., 2023; Eikenberry & Gumel, 2018; Mordecai et al., 2020; Peterson, 2009; Ryan et al., 2015). However, distilling global drivers at local scales has not yielded the same success due to the complex interactions between vectors, parasites, and humans that differ across the geographic range of malaria (Eikenberry & Gumel, 2018). Focusing on an area that is highly climatically suitable and endemic for malaria transmission (defined as an area where transmission occurs 10-12 months of the year; (Ryan et al., 2020)), we aim to assess how well coarse predictors of malaria transmission explain variation at a fine scale. Working in 40 villages in Côte d’Ivoire that participated in a randomized controlled trial of a household malaria intervention (screening and eave tubes (SET); (Sternberg et al., 2018; Sternberg et al., 2021)), we use data on *in situ* climate, malaria incidence, and entomological observations over a two-year period to study the effects of climate variation on malaria transmission. Specifically, we ask:

1. Which variables (climate or entomological metrics), and in what form (linear or unimodal, mechanistic or phenomenological) and time lags, best explain seasonal variation in malaria transmission?
2. What factors predict spatial variation of malaria transmission when climate is similar across sites?

## 2 Methods

### 2.1 Data collection

#### 2.1.1 Study location and design

Data were used from a previously published study (Sternberg et al., 2021) that aimed to assess the impact and cost-effectiveness of a novel malaria intervention in the Gbêkê region in central Côte d’Ivoire. Detailed study design and participant information can be found in Sternberg et al. (2018). In brief, epidemiological, entomological, and climate data were collected over a two-year period across 40 villages in the Gbêkê region. This region experiences high burdens of year-round malaria transmission, with peak transmission reported in the wet season (May-October). Twenty of the 40 villages received an intervention which combined Eave Tubes, a novel method of delivering insecticide at household level, and mosquito-proofing within households through the addition of window screening. The combined intervention (referred to as Screening plus Eave Tubes, or SET) was designed to reduce malaria transmission by a) providing a physical barrier between mosquito vectors and humans and b) increasing mosquito mortality as they search for human hosts for blood feeding.

#### 2.1.2 Epidemiological variables

Within each village a cohort of approximately 50 children (0.5-10 years old) was randomly selected. At the initial enrolment visit, all children were cleared of active malaria parasite infections with a 3-day course of first-line antimalarial drug (artesunate-amodiaquine or artemether-lumefantrine).

Following this initial visit, children were routinely visited (once per month between November and April, and twice per month between May and October) by medical staff to test for symptomatic malaria infections. During visits, children were treated with antimalarials if they were symptomatic and tested positive for malaria using a rapid diagnostic test (SD Bioline Malaria Ag P.f/Pan;Standard Diagnostics; Seoul, South Korea). Malaria incidence was calculated at the village level as a monthly measure of the number of clinical malaria cases divided by the child-time at risk (see Sternberg et al. (2018) for full details).

#### 2.1.3 Climate variables

To test the effects of rainfall and temperature on malaria transmission, we collected data from several sources. First, hourly rainfall and outdoor temperature data were collected from a single weather station within the study area (7°53′58″ N, 5°3′33″ W, sourced from AfricaRica upon request), and provided a study-wide estimate of these conditions. Second, we were interested in indoor temperatures, because *An. gambiae* are thought to be largely endophilic and spend considerable time indoors (Paaijmans & Thomas, 2011). Hourly indoor temperatures were collected from a subset of 20 (of the total 40) villages. Indoor temperature data collection was intermittent and not conducted in all villages, so to generate a study-wide measure of indoor temperature we rounded collection time to the closest hour to pair with outdoor meteorological station temperatures, and then fit a linear relationship between indoor temperature and outdoor temperature. We allowed the outdoor to indoor relationship to vary by hour of the day and month of the year, and used this to predict a study-wide indoor temperature estimate for each hour, resulting in a single time series of indoor temperature used across all villages.

#### 2.1.4 Entomological variables

Mosquitoes were trapped using human landing catches undertaken indoors and outdoors in four randomly selected houses per village every two months. Entomological inoculation rate (EIR; the number of infectious bites per person per year) was calculated every two months in each village. EIR was calculated as follows:

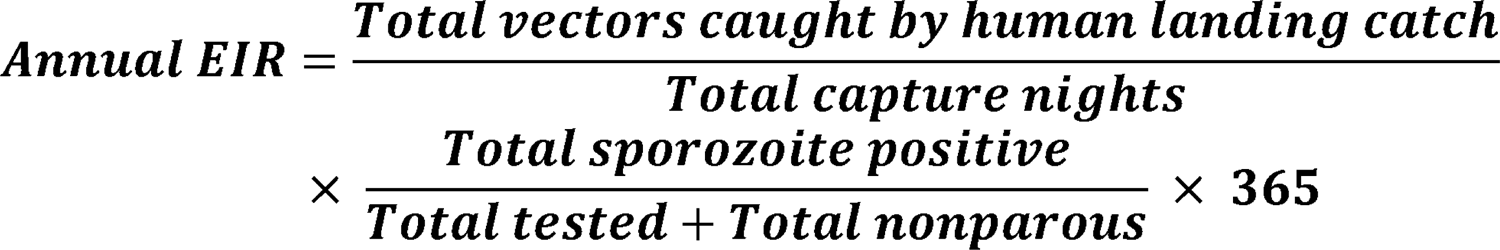

## 3 Data analysis

We conducted two main analyses to investigate the predictors of malaria incidence in the study region. The first analysis focused on understanding which variables best predicted variation over time across all villages (‘temporal analysis’) and the second aimed to explain spatial variation between villages (‘spatial analysis’).

### 3.1 Temporal analysis

To understand temporal variation in malaria incidence, we modeled climatic and entomological variables separately and quantified the variation explained by each variable. Panel regressions were used to isolate climate effects on malaria incidence from spatial village-level variation. Specifically, we used a Poisson panel regression with a fixed effect, or separate intercept, for village and a time trend (to account for the observed trend in malaria over time), using village-month observations of malaria incidence rates as the outcome. This method removes cross-sectional variation between villages, leaving behind what we call “temporal variation”, and identifies predictors of malaria incidence rate by comparing each village to itself over time (after controlling for the overall time trend shared across all villages). Unless otherwise specified, all models include village fixed effects and a time trend.

To compare between predictors of malaria transmission, we needed a way to standardize the measure of model performance (squared correlation between predicted and observed) given that data were collected at different frequencies and time periods. This was done first by estimating a lower (null model) and upper bound (flexible seasonal model) for model performance. For each predictor, performance was compared within these bounds. The null model was estimated as a village fixed effect with a time trend. The flexible seasonal model estimated a coefficient for each month of the year with the same village fixed effects and time trend, which we viewed as an upper bound on system predictability because it is allowed to fit a unique coefficient for each month, which broadly corresponds to seasonality. In this way, predictive models that had a squared-correlation similar to that of the null were considered to be low performing, and those performing as well as the flexible seasonal model are considered to be high performing.

In total, we consider three climatic variables (indoor temperature, outdoor temperature, and precipitation) and two entomological variables (outdoor EIR, indoor EIR). For each of these, we modeled different functional forms and lags (Table 1) to account for potential nonlinear relationships, and delays between changes in climatic conditions (or entomological conditions), changes in malaria transmission (which we cannot directly observe), and ultimate detection of clinical malaria infections in monthly visits. For both indoor and outdoor temperature, we model linear, quadratic, and *a priori*-informed R_0_ relationships. This R_0_ relationship is derived from laboratory studies that estimate mosquito life history and transmission traits at different temperatures, and provides an estimated relationship between constant temperatures and relative transmission rates into which we can plug the different temperature values (Villena et al., 2022). To account for the ways in which mosquitoes are exposed to indoor and outdoor temperatures, we also calculated a version of the R_0_ model that combines both temperature sources (which we refer to as combined R_0_), where outdoor temperature was used for larval traits and indoor temperature was used for adult traits associated with transmission. For precipitation, we fit models relating malaria incidence rates to linear and quadratic monthly precipitation. Finally, for indoor and outdoor EIR we use a linear relationship. Notably, while indoor temperature, outdoor temperature, and precipitation are study-wide predictors (i.e., a single time series for the study period), EIR is village specific. For all nonlinear relationships between climatic and entomological variables, we calculate the nonlinearities at the smallest available resolution (hourly for temperature and precipitation) before averaging to the month.

**Table 1.** Variables and their relative forms and lags included in the panel regression analysis.

| Variable | Data collection location | Functional forms | Lags |
| --- | --- | --- | --- |
| Temperature | Outdoor, indoor | Linear, quadratic, relative R0 (outdoor, indoor, and combined indoor and outdoor) | 0, 1, 2 months |
| Precipitation | Outdoor | Linear, quadratic | 0, 1, 2 months |
| EIR | Outdoor, indoor | Linear | 0, 1, 2 months |

### 3.2 Spatial analysis

To study the predictors of spatial variation in malaria incidence rate, we used the village mean malaria incidence rate over the study period as the response and considered land use, RCT treatment level, and entomological variables as predictors. Specifically, we used remotely sensed measures of the percentage of land cover composed of urban/built up, shrubs, cultivated area, open forest, or closed forest in a 1 km buffer around the village location from the Steinberg et al 2018 dataset; the percentage of houses in the village receiving the SET intervention; and the averages over the study period of indoor vector abundance, percentage of indoor vectors that were *Anopheles gambiae*, and indoor EIR. Remotely sensed land cover variables are from the discrete classifications in the Copernicus Global Land Cover Layers in 2018 (Buchhorn et al., 2020), which we grouped into the 5 categories noted above.

To contextualize the estimated impacts of each of the covariates on malaria incidence rates, we used the 5th and 95th percentiles of each covariate, and calculated the change in malaria associated with a covariate increasing from its 5th to 95th percentile.

## 4 Results

### 4.1 Climate and entomological predictors of malaria seasonality

Malaria incidence per child per year followed a clear seasonal pattern (Figure 1a), with peaks in incidence occurring between August and November, and lowest incidence between January and April, in both treated and untreated villages. Malaria incidence was higher in villages that did not receive the SET treatment (Figure 1a), as shown previously (Sternberg et al., 2021). EIR varied sporadically over the study period, although as with malaria incidence, it was higher on average in control than treated villages (Figure 1a). Temperature (indoor and outdoor) and rainfall also had consistent seasonal variation, and indoor temperatures were consistently warmer than outdoor temperatures (Figure 1a). As outdoor temperatures varied around the presumed optimal temperature for malaria transmission (25°C, Mordecai et al. (2013)), estimated relative R_0_ as a function of outdoor temperature was relatively high for an extended period of the year (March to October) while indoor temperatures—which were consistently above the optimal temperature—lead to an estimated relative R_0_ with two sharper peaks in the cooler times of the year (Figure 1a). The combined R_0_, which used outdoor temperatures for larval traits and indoor temperatures for adult traits, had a smoother seasonal pattern with a single, narrow peak in July to September that matches the seasonality of malaria rate and indoor EIR (Figure 1a).

**Figure 1:**
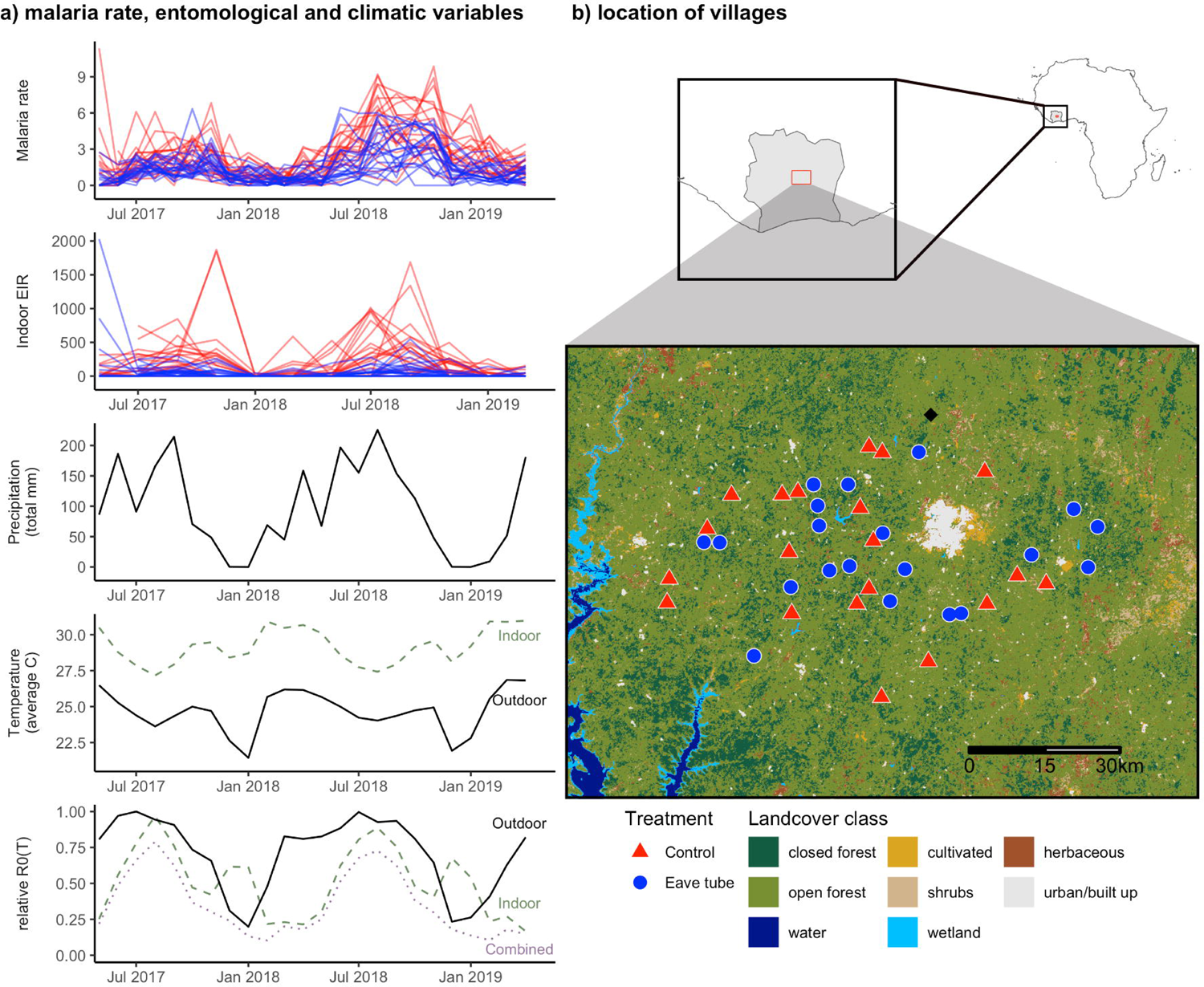
**a)** Malaria incidence rate (incidence per child per year) and indoor EIR (infectious bites per person per year) in control (red) and eave tube treated (blue) villages over the study period. Precipitation (monthly total, mm) and temperature (monthly average, °C) from a local meteorological station. Dashed horizontal line indicates the predicted temperature of peak R0 from mechanistic models (Mordecai et al., 2013). Bottom panel shows relative R0(T), calculated as the average of daily R0(T) over the month (Mordecai et al., 2013). **b)** Location of control (red triangle) and treated (blue circle) villages, location of meteorological station (black diamond), and land cover in the area.

Climate variables were available every month, so models predicted monthly malaria incidence rates with predictors lagged at 0, 1 and 2 months. Among these models, the lower-bound null model explained 23% of malaria variation (measured by squared correlation between predicted and observed of the Poisson regression), while the flexible seasonal model explained a total of 68% of malaria variation (Figure 2). Generally, the quadratic models for both indoor and outdoor temperatures (explaining 63% and 65% of malaria variation, respectively) performed much better than linear temperature models (55% and 26% of malaria variation), while the R_0_ model that combined indoor and outdoor temperature performed slightly better than outdoor or indoor R_0_ temperature alone (65% vs 60% and 55%, respectively; Figure 2). For rainfall, both linear and quadratic models performed well (62% and 64%, respectively; Figure 2). While the best performing climatic variables approached the performance of the flexible model, with both quadratic temperature and rainfall explaining the greatest variation in malaria transmission (Figure 2), it is not possible to identify which may be the main seasonal driver because these variables are highly correlated.

**Figure 2:**
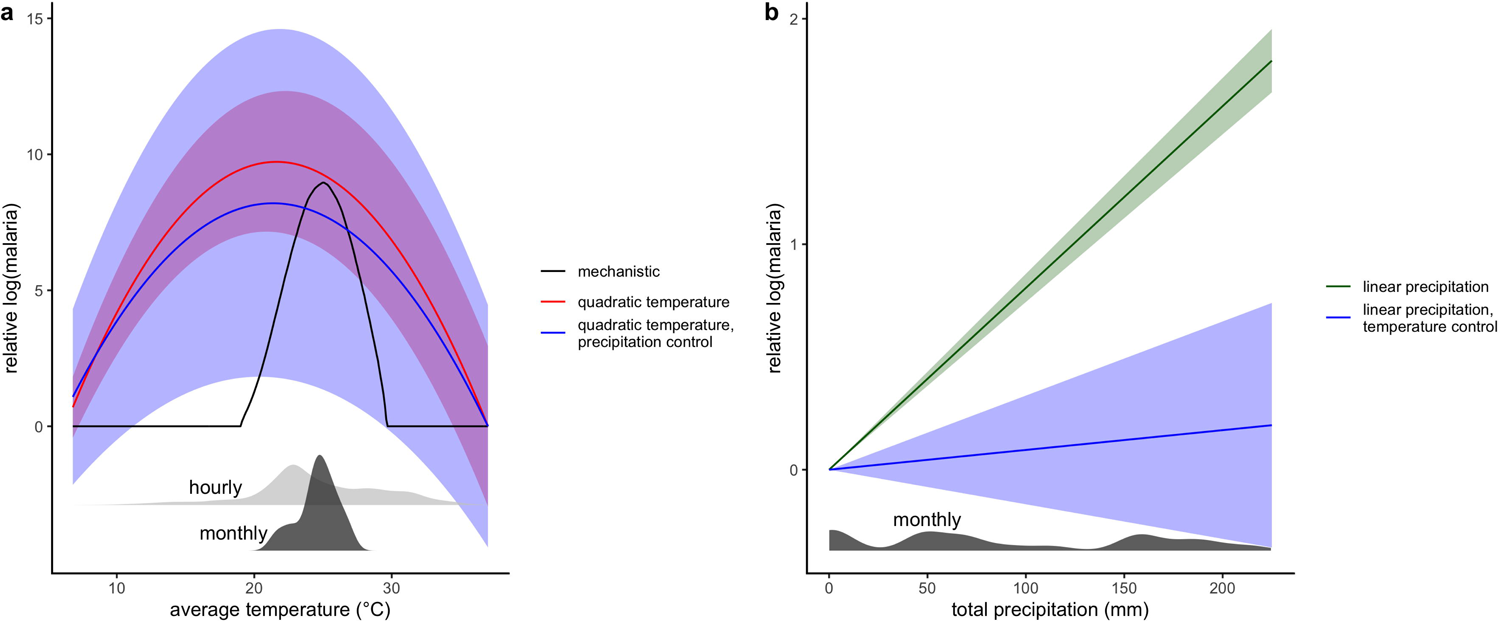
Nonlinear temperature (quadratic and R_0_) and rainfall explain substantial variation in malaria incidence. Relative performance of models, measured by relative squared correlation from models, where the minimum of 0 is squared correlation equivalent to a baseline model with only village fixed effects (FEs) and a time trend (vertical lines on the left) and the maximum of 1 is relative to model with village FEs, time trend and flexible seasonal (month FE; vertical lines on the right). Models shown below the dashed line are on the sample that has data on EIR (i.e., only bi-monthly) and includes only current and 2-month lagged predictor variables for consistency with the availability of EIR data

Entomological predictor variables were only available every other month, so models predicted bi-monthly malaria incidence rates, and predictors were lagged at 0 and 2 months. For these models, the null model explained 26% of malaria variation, and the flexible seasonal model explained 68% of malaria variation (Figure 2). We found that both indoor and outdoor EIR performed lower than expected (44% and 33%; Figure 2) given that it has a tight relationship to malaria transmission in the literature (Kelly-Hope & McKenzie, 2009). To ensure this was not due to the bimonthly nature of the predictive models, we also fit bimonthly models with quadratic outdoor temperature and combined R_0_ temperature (using the same 0 and 2 months lags) and find that they outperform the EIR models (65% and 60%, respectively) and are much closer to the predictive power of the flexible seasonal model (Figure 2).

### 4.2 Theoretical vs. observed optimal temperature for malaria transmission

We tested the hypothesis that a temperature-dependent R_0_ model based on laboratory studies, which peaks at 25°C, would predict variation in malaria incidence rates, study-wide. We found some support for this hypothesis: our combined indoor and outdoor temperature-dependent R_0_ model explained 65% of the variation in malaria incidence rate, nearly matching the performance of the more flexible quadratic temperature model that explained 68% of variation (Figure 2). The estimated peak of the fitted quadratic outdoor temperature model occurred at 21.6°C, a few degrees cooler than the a priori predicted thermal optimum temperature of 25°C (Figure 3). However, indoor temperatures were consistently higher than outdoor temperatures (Figure 1a), and *Anopheles gambiae* mosquitoes spend a large portion of their adult lifecycle, indoors, which may explain this difference. This is also consistent with the improved performance of combined temperature R_0_ over indoor or outdoor R_0_, although the difference in performance of outdoor and combined R_0_ was small (Figure 2). In sum, temperature had a hump-shaped relationship with malaria incidence rate, as theoretically expected based on ectotherm physiology and laboratory thermal performance measurements (Johnson et al., 2015; Mordecai et al., 2013; Villena et al., 2022). While this temperature-dependent model had previously been tested against field data on spatial variation in malaria transmission (Johnson et al., 2015; Mordecai et al., 2013; Villena et al., 2022), this is important independent field evidence that it can potentially explain part of the observed variation in malaria over time within a small geographic area with high transmission rates.

**Figure 3:**
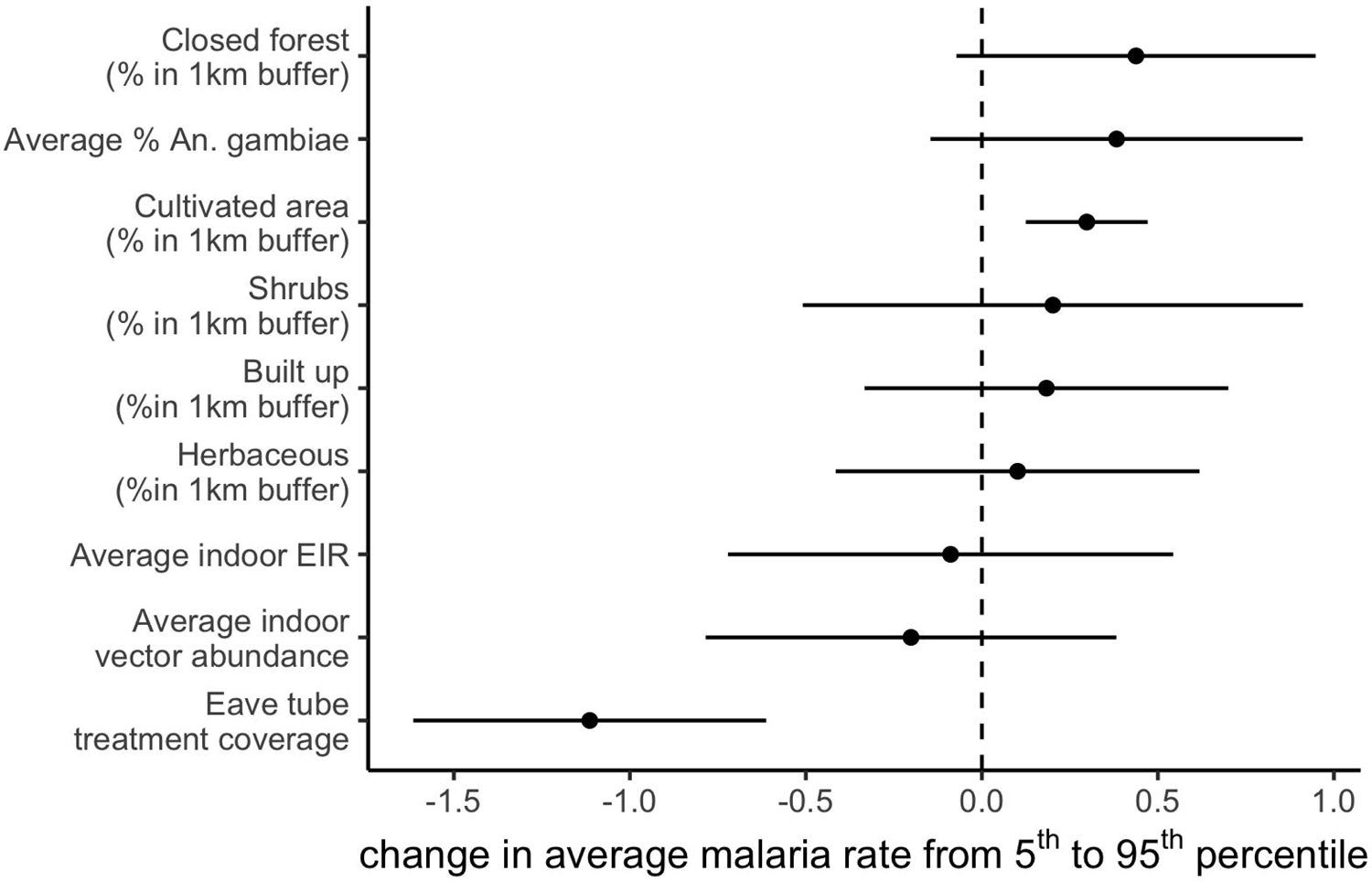
Estimated (a) temperature and (b) precipitation responses from panel regressions. **a)** Temperature responses use quadratic relationship from outdoor temperature in models with only temperature (red) and with linear precipitation control (blue), compared to mechanistic R_0_(T) curve (black). Solid line shows central estimate and shaded areas show 95% confidence intervals. Density plots show the distribution of monthly average (dark grey) and hourly (light grey) outdoor temperatures. b) Precipitation responses similarly are from models with just linear temperature (green) and with quadratic temperature control (blue).

### 4.3 Effects of rainfall on malaria transmission

Unlike temperature, we did not have precise a *priori* predictions for how rainfall would affect malaria incidence rate. We did, however, expect that transmission would be most strongly associated with rainfall at lags of 0-2 months, given the time needed after rainfall for mosquitoes to hatch, develop to adulthood, bite, acquire parasites, become infectious, and transmit onward. We found that both linear and quadratic relationships between rainfall (lagged 0 - 2 months) were predictive of malaria incidence rates, with little improvement from the additional flexibility of the quadratic model (62% vs 64% R^2^; Figure 2), in contrast to temperature. This relationship showed increasing malaria with higher monthly rainfall, but, after additionally controlling for quadratic temperature, this association was much weaker, indicating the highly correlated nature of the seasonal signals of temperature and precipitation (Figure 3).

### 4.4 Spatial variation in malaria transmission

Study-wide malaria followed a consistent seasonal pattern over time but malaria incidence rates varied substantially between villages, particularly during the second transmission season (2018), which coincides with reducing efficacy in insecticide-treated bed nets distributed at the start of the trial (Sternberg et al., 2021). We were interested in understanding what factors, in addition to the SET treatment, explained village-level differences in malaria incidence rate in this geographically and climatically similar group of villages. We used remotely sensed vegetation cover and field-measured entomological variables as predictors to explain variation in the village-level average malaria incidence rate over the study period. Although microclimatic, demographic, and socioeconomic factors likely also affect malaria incidence rates, we did not have access to data on these variables. We found that in addition to SET treatment reducing malaria incidence rate (as shown previously; Sternberg et al., 2021), the percentage of cultivated land area at 1 km scale was associated with increased malaria incidence rate (Figure 4). Entomological variables, averaged by village over the entire study period (two years), were not significantly associated with average malaria incidence rates among villages. This indicates that, for example, villages with consistently higher malaria incidence rates did not have consistently higher entomological indices (including average indoor vector abundance, indoor EIR, and percentage of vectors that are *An. gambiae*).

**Figure 4:**
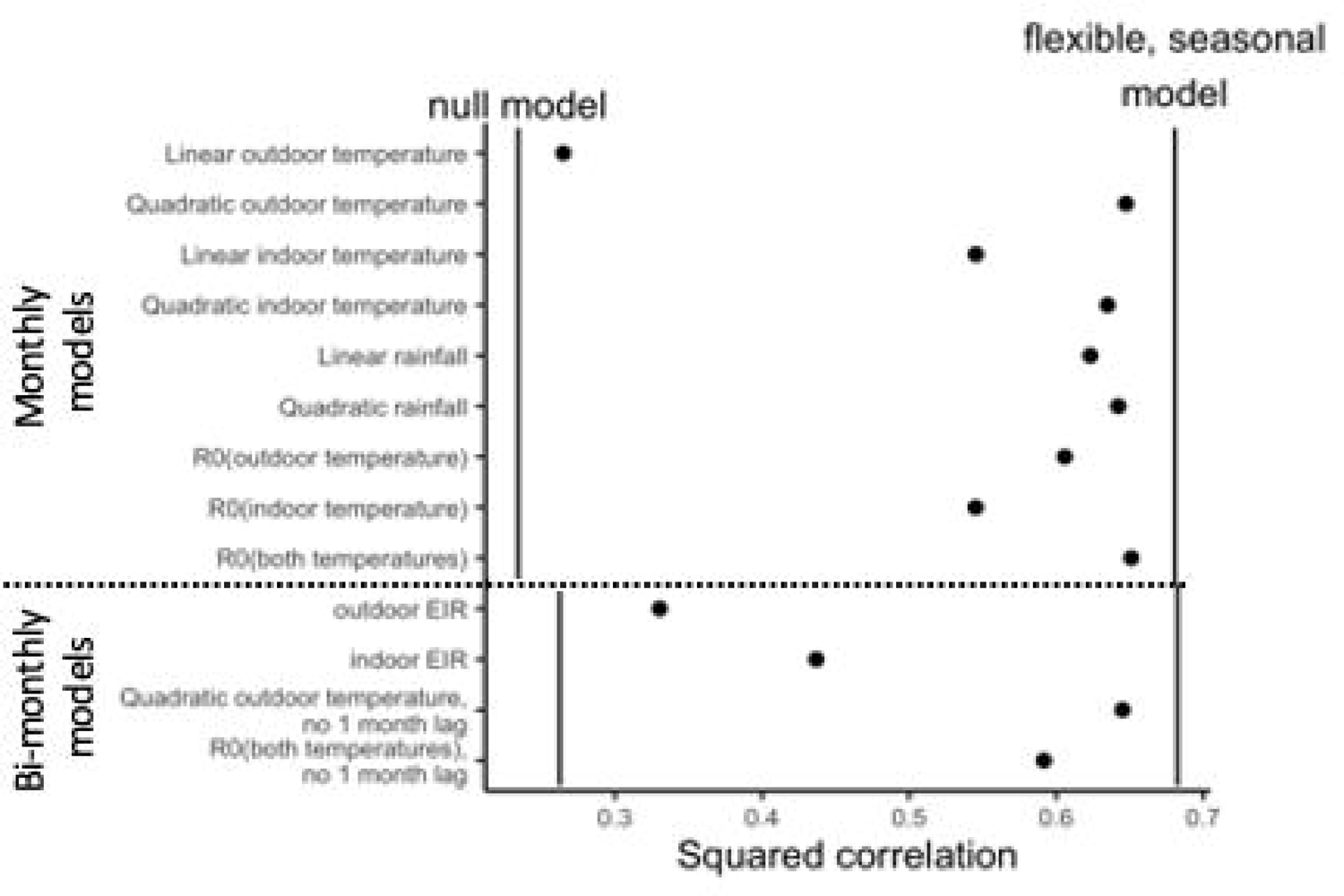
Cultivated area increases malaria incidence rate while SET treatment decreases it. Estimated change on average malaria incidence rate from changing a covariate from the 5th to 95th percentile of its distribution. Points indicate central estimates and error bars are 95% confidence intervals. Estimates are from a linear regression of average malaria incidence on covariates.

## 5 Discussion

The transmission of vector-borne diseases depends on a suite of environmental, climatic, and social factors. The importance of these variables, and their mechanistic links with transmission vary across pathogens, vectors, locations, and scales. Climate and malaria are one of the best studied interactions, given that malaria i) causes one of the highest burdens of all vector-borne diseases globally and ii) is a historically important disease which has been investigated for over a century. In recent decades we have advanced our understanding of relationships between climate and malaria at large geographic scales, which is essential for predicting the distribution of malaria and shifting environmental suitability under climate change scenarios. For example, a recent study using panel regression methods applied across sub-Saharan Africa from 1900-2015 found that temperature had nonlinear effects on *P. falciparum* rates in 2-10 year-olds, which peaked at 25°C, and that historical climate change has already led to geographical shifts in malaria burdens from historically warm West and Central Africa to historically cooler regions of East and Southern Africa (Carlson et al., 2023). Yet, in localized areas that are highly suitable for transmission and experience high burdens of malaria, it remains unclear how important climate is for predicting malaria incidence.

Here we modeled multiple climate variables, in different functional forms and lags, across 40 villages in a holoendemic area of malaria transmission. When assessing the performance of temperature as a predictor of malaria, we find strong support for the emerging consensus that effects of temperature are nonlinear. In our study, nonlinear temperature performed almost as well as the upper bound measure (a flexible seasonal model), while linear temperature performed much closer to the lower bound measure (a null model). Over the last decade, a growing body of literature has provided empirical evidence for the nonlinear effects of temperature on transmission, which are attributed to the thermal performance of all vector and parasite life history traits (Carlson et al., 2023; Johnson et al., 2015; Mordecai et al., 2013; Peterson, 2009; Shah et al., 2019; Villena et al., 2022; Yamana & Eltahir, 2013). Our findings add further support to this and highlight that warmer temperatures do not always lead to more malaria. In addition to the linear and quadratic temperature models, we included a mechanistic R_0_ of temperature model based on outdoor temperatures, indoor temperatures, and one that integrated both indoor and outdoor temperatures specific to malaria vector behavior and biology. Of these three, the combined indoor and outdoor temperature model performed best. In fact, it performed as well as the upper bound (flexible seasonal) model, and slightly better than the quadratic temperature model (Figure 2). This is a novel finding because few models combine indoor and outdoor temperature data or acknowledge that larvae and adults might experience different temperature. In general, a mechanistic R_0_ model can be more informative than phenomenological nonlinear models alone, particularly when correlated climate variables makes it difficult to assess causation, because they integrate components of vector biology and transmission that are specific to transmission risk. However, in comparison to a phenomenological nonlinear model they can be more complex to develop and require additional data on vector biology to validate findings in a given location.

Due to the strong seasonal correlation between temperature and rainfall, we found that a model incorporating rainfall performed as well as the nonlinear temperature models, and the difference between linear and nonlinear rainfall patterns was marginal. The quantitative effects of rainfall on malaria are less broadly understood compared to those of temperature. In some studies, an increase in rainfall has been associated with more malaria due to the increased availability of vector breeding sites, yet too much rainfall has also been associated with a reduction in malaria because it can flush larval habitats (Asare et al., 2016; Ratti et al., 2022). The functional form relating rainfall to malaria may vary across settings that differ in climate, hydrology, and land use. Our findings underscore two considerations for using rainfall as a local predictor of malaria. First, the high performance of both functional forms emphasizes the need for finer scale assessments that can account for localized variation in hydrology. Our study encompassed 40 villages which may differ slightly in hydrological profiles, but we were limited by a single measure of rainfall across the study area. Second, because both temperature and rainfall vary seasonally in a pattern that closely matched malaria transmission, it is not possible to disentangle which of these variables are more important for driving transmission using the information in these models alone.

The randomized controlled trial (RCT) underpinning this study provides an unprecedented opportunity to connect climate, entomological variables, and malaria incidence measured consistently across a large set of 40 villages over a 2-year period. Specifically, entomological variables like mosquito abundance, entomological inoculation rate (EIR), and percentage of vectors that are *An. gambiae* are often measured as potential indicators of malaria transmission (Kelly-Hope & McKenzie, 2009). However, we found that for variation both over time and among villages, entomological predictors were not strongly associated with malaria incidence rate, and that climatic variables are better predictors of variation in transmission. This may be because mosquito sampling methods are inherently noisy and because entomological variables were only measured every other month, and were averaged within villages in all analyses. It is possible that more finely resolved entomological data could have greater predictive power. Still, given that these data are costly to collect and that climatic variables explained more variation in malaria incidence rate within villages over time (they were not measured separately among villages), this work suggests that mechanistic models of the effects of climate on malaria transmission could be useful tools for predicting variation in transmission over time and space. Entomological indices may be valuable as a proximate measure of the impact of vector control interventions and for identifying particular households that are at higher risk.

The focal villages were selected for inclusion in the RCT based on their socio-ecological similarity and geographic proximity; still, they varied substantially in malaria incidence, from 0-11.35 malaria cases per child, and much of this variation was not explained by the SET treatment. We found that villages with more surrounding cultivated land cover had higher malaria incidence rates, but that most of the variation among villages remained unexplained by our land cover, entomological, and SET treatment variables. This suggests that unmeasured factors like demography, mobility, socioeconomic conditions, built infrastructure, and microclimate variation may mediate malaria transmission in this highly endemic setting: an important area for further study.

## 6 Conclusion

While recent decades have seen dramatic declines in transmission across the continent due to improvements in malaria control efforts (Bhatt et al., 2015), malaria remains paradoxically difficult to control in many endemic areas, and progress toward malaria elimination has recently stalled (World Health Organization, 2022). For example, despite this RCT testing for and clearing parasites prior to and throughout the study, providing new insecticide-treated bednets, and providing a novel household modification to reduce mosquito entrance and survival (SET), malaria case incidence was 2.29 per child-year in control villages and 1.43 per child-year in treated villages during the 2-year intervention period. Understanding the burden of malaria and how it responds to climate is therefore critical for predicting malaria incidence over space and time and for supporting targeted interventions. Despite the complexity of the malaria transmission system, we found that up to 68% of within-village variation over time is predictable based on seasonality of rainfall and temperature. Accounting for nonlinear effects of temperature on malaria transmission is critical as linear models of temperature performed poorly at explaining malaria incidence rates (Figure 2). This provides further empirical support for an emerging understanding that climate warming is likely to drive geographic and seasonal shifts rather than across-the-board increases (Carlson et al., 2023; Mordecai et al., 2020; Ryan et al., 2015). Preparation for these climate change-driven shifts is a major objective for public health and sustainable development in the 21st century.

## 7 Conflict of Interest

The authors declare that the research was conducted in the absence of any commercial or financial relationships that could be construed as a potential conflict of interest.

## 8 Author Contributions

EBS, MLC and EAM planned the methods and interpreted the results, MLC carried out the modelling, and EBS took the lead in writing the manuscript. MBT, JC, EDS conceived the presented idea, collated data and contributed to interpretation of results. AAK, RN, RZQ, WAO, LPAA, SB collected primary data. All authors helped shape the research and approved of the manuscript.

## 9 Funding

E.A.M. and E.B.S. were supported by the National Institutes of Health (grant no. R35GM133439) and E.A.M. was supported by the National Science Foundation and the Fogarty International Center (grant no. DEB-2011147) E.A.M. was additionally supported by the National Institute of Allergy and Infectious Diseases (grant nos. R01AI168097 and R01AI102918) and by seed grants from the Stanford Woods Institute for the Environment, King Center on Global Development, Center for Innovation in Global Health and Terman Award. M.L.C was supported by the Illich-Sadowsky Fellowship through the Stanford Interdisciplinary Graduate Fellowship program at Stanford University and an Environmental Fellowship at the Harvard University Center for the Environment. The empirical data used in this study derive from a project supported by the Bill & Melinda Gates Foundation (OPP1131603).

## Data Availability

The datasets analysed for this study are available upon request. Code to reproduce all results and figures in the manuscript will be made available upon publication at https://github.com/marissachilds/ivory-coast-malaria-predictability.

## Acknowledgments

We thank AfricaRice for providing hourly climate data. We also acknowledge Joelle I. Rosser and the Mordecai lab for constructive feedback throughout the development of this study.

## 10 Supplementary Material

Supplementary Material should be uploaded separately on submission, if there are Supplementary Figures, please include the caption in the same file as the figure. Supplementary Material templates can be found in the Frontiers Word Templates file. Please see the Supplementary Material section of the Author guidelines for details on the different file types accepted.

## Notes

### Competing Interest Statement

The authors have declared no competing interest.

### Author Declarations

The case data involving humans were approved by Cote d'Ivoire Ministry of Health ethics committee (039/MSLS/CNER-dkn),the Pennsylvania State University's Human Research Protection Program under the Office for Research Protections (STUDY00003899 and STUDY00004815), and the London School of Hygiene & Tropical Medicine ethical review board (11223). The studies were conducted in accordance with the local legislation and institutional requirements. Written informed consent for participation in this study was provided by the participants' legal guardians/next of kin.

